# Augmenting contact matrices with time-use data for fine-grained intervention modelling of disease dynamics: A modelling analysis

**DOI:** 10.1101/2020.06.03.20067793

**Authors:** Edwin van Leeuwen, PHE Joint modelling group, Frank Sandmann

## Abstract

**Background:** Social distancing is an important public health intervention to reduce or interrupt the sustained community transmission of emerging infectious pathogens, such as SARS-CoV-2 during the coronavirus disease 2019 (COVID-19) pandemic. We aimed to explore the impact on the epidemic curve of fewer contacts when individuals reduce the time they spend on selected daily activities.

**Methods:** We combined the large-scale empirical data of a social contact survey and a time-use survey to estimate contact matrices by age group (0-15, 16-24, 25-44, 45-64, 65+) and daily activity (work, schooling, transportation, and four leisure activities: social visits, bar/cafe/restaurant visits, park visits, and non-essential shopping). We assumed that reductions in time are proportional to reductions in contacts. The derived matrices were then applied in an age-structured dynamic-transmission model of COVID-19 to explore the effects.

**Findings:** The relative reductions in the derived contact matrices were highest when closing schools (in ages 0-14 years), workplaces (15-64 years), and stopping social visits (65+ years). For COVID-19, the closure of workplaces, schools, and stopping social visits had the largest impact on reducing the epidemic curve and delaying its peak, while the predicted impact of fewer contacts in parks, bars/cafes/restaurants, and non-essential shopping were minimal.

**Interpretation:** We successfully augmented contact matrices with time-use data to predict the highest impact of social distancing measures from reduced contacts when spending less time at work, school, and on social visits. Although the predicted impact from other leisure activities with potential for close physical contact were minimal, changes in mixing patterns and time-use immediately after re-allowing social activities may pose increased short-term transmission risks, especially in potentially crowded environments indoors.

**Research in context:** *Evidence before this study:* We searched PubMed for mathematical models using social contact matrices and time-use data to explore the impact of reduced social contacts as seen from social distancing measures adopted during the coronavirus disease 2019 (COVID-19) pandemic with the search string ((social OR physical) AND distancing) OR (contact* OR (contact matri*)) AND (time-use) AND (model OR models OR modeling OR modelling) from inception to May 06, 2020, with no language restrictions. We found several studies that used time-use data to re-create contact matrices based on time spent in similar locations or to calculate the length of exposure. We identified no study that augmented social contact matrices with time-use data to estimate the impact on transmission dynamics of reducing selected social activities and lifting these restrictions again, as seen during the COVID-19 pandemic.

*Added value of this study:* Our study combines the empirical data of two large-scale, representative surveys to derive social contact matrices that enrich the frequency of contacts with the duration of exposure for selected social activities, which allows for more fine-grained mixing patterns and infectious disease modelling. We successfully applied the resulting matrices to estimate reductions in contacts from social distancing measures such as adopted during the COVID-19 pandemic, as well as the effect on the epidemic curve from increased social contacts when lifting such restrictions again.

*Implications of all the available evidence:* Social distancing measures are an important public health intervention to limit the close-contact transmission of emerging infectious pathogens by reducing the social mixing of individuals. Our model findings suggest a higher fraction of close-contact transmission occurs at work, schools, and social visits than from visits to parks, bars/cafes/restaurants, and non-essential shopping. The minimal predicted impact is suggestive of lifting the restrictions on certain activities and excluding them from the list of social distancing measures, unless required to maintain sufficient healthcare capacity. However, potential replacement effects of activities and in mixing patterns remain unclear, particularly immediately after re-allowing social activities again.

## Introduction

Social contacts between individuals in close physical proximity drive the transmission dynamics of many respiratory infections such as influenza virus and coronavirus [1,2]. In community outbreaks of emerging infectious diseases, such as with the coronavirus disease 2019 (COVID-19) pandemic, social distancing measures are an important public health intervention. (Note that we use the term “social distancing” in line with convention; however, it aims at avoiding physical contacts only, while other non-physical interactions via information and communication technologies are encouraged [3].) Measures of social distancing aim to reduce or interrupt the spread between individuals of unknown infection status by reducing the mixing of populations [3]. Individuals are requested to make fewer and shorter contacts by changing their daily activities and the locations at which they spend their time.

Contact matrices are typically used when evaluating such public health interventions as they account for the heterogeneity in mixing of individuals in a *n* × *n* matrix, with *n* equal to the number of different age groups [4]. Empirical contact matrices are derived from contact surveys, which collect data on the frequency and location of social interactions between individuals by age, with some collecting limited information on the duration too [5]. However, these matrices often lack detailed information on the time individuals spend on daily activities, and the COVID-19 pandemic highlighted the need for a more fine-grained modelling of contact patterns [6]. In contrast to contact surveys, time-use surveys collect information on how much time individuals spend per day on a wide range of social activities, at what location, and with some limited information on with whom [7]. Previous studies have used time use data to re-create contact matrices based on time spent in similar locations [8,9] or used time-use data to calculate the length of exposure [10]. This study combined both data sets to estimate number of contacts per activity, and we applied the resulting matrices to estimate reductions in contacts from social distancing measures as adopted during the COVID-19 pandemic.

This study aimed to augment existing social contact matrices with time-use data to estimate the effect of social distancing measures for the number of contacts per day when reducing the time spend on selected daily activities (some of which are considered to be non-essential). The derived contact matrices were based on the empirical data of two large-scale population-wide surveys, allowing for more fine-grained intervention modelling. Afterwards, we applied the derived contact matrices in a dynamic-transmission model to explore the impact on the COVID-19 pandemic, which was characterised by many countries with sustained community transmission adopting wide scale social distancing measures at population-level [3,11,12].

## Methods

### Datasets

We used two large-scale datasets that included information on the age-mixing patterns of individual contacts by location and daily activity. The contact matrices were obtained from the POLYMOD study [13], which was a nationally-representative social contact survey that collected data on the frequency, duration, type (physical or non-physical) and location (home, work, school, leisure, transport, or other) of social interactions in 7,290 individuals for each of their 97,904 contacts in eight European countries in 2005-2006. Participants filled out a diary for 24 hours on a randomly selected weekday or weekend day, recording physical skin-to-skin contacts (e.g. a kiss or a handshake) versus non-physical contacts (a two-way conversation of more than two words without skin-to-skin contact). The age-mixing patterns were similar across countries, and revealed a strong assortativeness, i.e. individuals spend more time with others of their same age [13].

Individual time-use data were obtained from the United Kingdom Time Use Survey (UKTUS) [14], which was a nationally-representative survey that collected data on the frequency, duration, and location of a wide range of daily activities in 16,550 diary entries of 9,388 individuals aged 8+ years from the UK in 2014-2015, and whether activities were spent alone or with others. Participants were asked to complete diaries for 24 hours on two randomly selected days (one on a weekday, one on a weekend day), recording sequences of activities at intervals of 10 minutes [14]. All activities in the time use survey were grouped according to codes. These codes were then associated with certain POLYMOD locations and activities using Table S1 in the supplementary materials.

### Contact by location and activity

Contact studies such as POLYMOD measure the number of contacts (*k_il,j_*) by age group (*i*) and location of the participant (*l*) and the contacts age group (*j*). Time-use surveys such as the UKTUS measure the time (*t_ial_*) spent by age group (*i*), activity (*a*) and location (*l*). This makes it possible to estimate the contact by activity and location as follows:

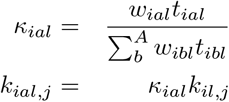

with *κ_ial_* the activity weight for age group *i*, activity *a* at location *l*, *t_ial_* is the average time spent by an individual, *A* is the set of activities and *w_ial_* is an activity specific weight, which reflects the relative number of people met during this activity compared to other activities at the same location. Most weights will be kept equal to 1 unless specified otherwise.

We considered five age groups (0-15, 16-24, 25-44, 45-64, 65+) as well as the six locations used in POLYMOD (home, work, school, leisure, transport, or other). The selected daily activities with potential for non-essential close physical contact were work, schools (i.e., educational settings of schools or universities, hereafter just called schools), transportation, social visits (i.e., visiting other people’s home and receiving visitors in one’s own home as part of celebrations and social life), bar/cafe/restaurant visits, park visits, and non-essential shopping. For a full list of the codes used to map activities to locations see Table S1 in the supplementary materials.

When an individual changes the relative time spent on an activity (*r_ial_*), while everyone else’s behaviour stays the same, the weight changes relative to the change in time 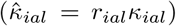. Of course, in reality the behaviour of the contact will also have changed and this also influences the probability of a contact during that activity and at that location. This is further complicated by the fact that the activity and location of the contact can be distinct from the activity and location of the participant. For example, in a restaurant the staff’s location and activity would be work, while the patron’s location and activity would be leisure and restaurant. The changes in the contact’s behaviour at the participant’s activity (*a*) and location (*l*) 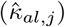, therefore, could be distinct from changes in the participant’s behaviour 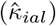. Still, this behaviour is likely to be proportional, e.g. if fewer people go to a restaurant then the restaurant will also reduce its number of staff. Furthermore, for most contacts the activities and locations will likely be the same. For this study we thus assume that the contact’s change in behaviour is in line with changes of behaviour relevant for that activity, location and the contact’s age group (i.e. 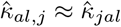).

Under the assumptions above, when an encounter happens at random then the probability of the encounter occurring is relative to the reduction of activity by both individuals as follows: 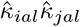. If the encounter is initiated by either individual, and we assume that either contact is 50 percent likely to be the initiator, then the probability is: 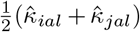. Therefore, we also need to know the fraction of contacts that were initiated by one of the individuals compared to being at random (*q_al_*). The new probability following changed activity is, therefore, equal to

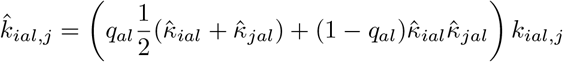

Using the estimated change in the number of contacts the contact matrix can then be calculated as follows: 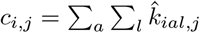.

As less time is spent on certain activities, more time will be spent doing other activities. For this study we assume that the replacement activities mostly do not increase the number of contacts, partly because they would be spent alone (exercise) or at home/with members of the household. While extra time spent at home would increase the number of contact events, most of those encounters will be with other members of the household and therefore the number of unique contacts will not increase linearly. To reflect this distinction, we assumed the number of contacts in the household is independent of the (extra) time spent there.

### Epidemiological model

The potential effect of the reduced contacts following social distancing interventions was explored by using the derived contact matrices as part of an epidemiological dynamic-transmission model. Epidemiological disease dynamics of COVID-19 were modelled using an age stratified SEEIIR model, which was based on models used in influenza modelling [15,16] and the latent and infectious periods were assumed to be 4.6 and 5 days long, respectively, in line with estimates for SARS-CoV-2 [17]. The basic reproductive number (*R*_0_) was assumed to be 2.8. This age-stratified model made it possible to estimate the effect of changes in the incidence when the time spent performing certain activities is being changed, and to explore the impact on specific at-risk individuals such as the elderly aged 65+ years.

We present the results of two main scenarios. First, to examine the contribution of different activities to transmission, we allowed all but one activity to continue. Second, allowing individual activities to resume again while all other activities continue to be reduced.

## Results

Table 1 shows the relative contribution of each activity to the contacts at the POLYMOD locations (calculated using 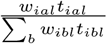). Social visits were the most important leisure activity regardless of age. Bars, cafes and restaurants are the second most important leisure activity except in children, where indoors exercise is more important. The results also showed that a large share of the time spend on transport is linked to school and work activities. Therefore, if school and work activities were reduced then this will also have a considerable impact on the contact pattern during transport.

**Table 1:** Relative weight of each activity on contacts by POLYMOD location and age group. Home alone and bed were weighted as zero to reflect that no (new) contacts occur when alone or in bed (for sleeping, not asleep, or sick).

| Location | Activity | [0,16) | [16,25) | [25,45) | [45,65) | [65,+) |
| --- | --- | --- | --- | --- | --- | --- |
| home |  | 0.980 | 0.976 | 0.980 | 0.978 | 0.965 |
|  | alone | 0.000 | 0.000 | 0.000 | 0.000 | 0.000 |
|  | sleep | 0.000 | 0.000 | 0.000 | 0.000 | 0.000 |
|  | visit | 0.020 | 0.024 | 0.020 | 0.022 | 0.035 |
| leisure | bars and restaurants | 0.042 | 0.134 | 0.209 | 0.229 | 0.196 |
|  | cultural | 0.063 | 0.059 | 0.073 | 0.080 | 0.112 |
|  | exercise | 0.146 | 0.129 | 0.136 | 0.144 | 0.130 |
|  | library | 0.001 | 0.002 | 0.004 | 0.003 | 0.006 |
|  | parks | 0.092 | 0.024 | 0.063 | 0.057 | 0.031 |
|  | visit | 0.655 | 0.652 | 0.515 | 0.486 | 0.526 |
| otherplace | holiday | 0.173 | 0.124 | 0.184 | 0.210 | 0.149 |
|  | shopping | 0.174 | 0.226 | 0.223 | 0.248 | 0.287 |
|  | shopping essential | 0.039 | 0.058 | 0.080 | 0.078 | 0.070 |
|  | sleep | 0.000 | 0.000 | 0.000 | 0.000 | 0.000 |
|  | unspecified | 0.613 | 0.592 | 0.513 | 0.464 | 0.494 |
| school | school | 1.000 | 1.000 | 1.000 | 1.000 | 1.000 |
| transport | cultural | 0.083 | 0.052 | 0.074 | 0.053 | 0.060 |
|  | exercise | 0.071 | 0.046 | 0.029 | 0.020 | 0.026 |
|  | holiday | 0.031 | 0.031 | 0.019 | 0.030 | 0.031 |
|  | parks | 0.034 | 0.016 | 0.023 | 0.021 | 0.035 |
|  | school | 0.220 | 0.131 | 0.053 | 0.019 | 0.008 |
|  | shopping | 0.076 | 0.088 | 0.102 | 0.110 | 0.166 |
|  | transport | 0.288 | 0.252 | 0.292 | 0.357 | 0.440 |
|  | visit | 0.180 | 0.199 | 0.145 | 0.127 | 0.156 |
|  | work | 0.017 | 0.184 | 0.265 | 0.263 | 0.077 |
| work | work | 1.000 | 1.000 | 1.000 | 1.000 | 1.000 |

Using the results from Table 1 it was possible to calculate the difference in the contact matrix if certain activities were reduced. Figure 1 highlights the impact of stopping these activities on the contact matrix. Closing schools, clearly had the highest relative effect on contacts in the young age groups (ages 0-24 years), but still had effect on the number of contacts in adults as well. Work had by far the highest effect in adults aged 16-64 years. If we focus on the elderly age group then reducing social visits had the highest impact on reduced contacts.

**Figure 1:**
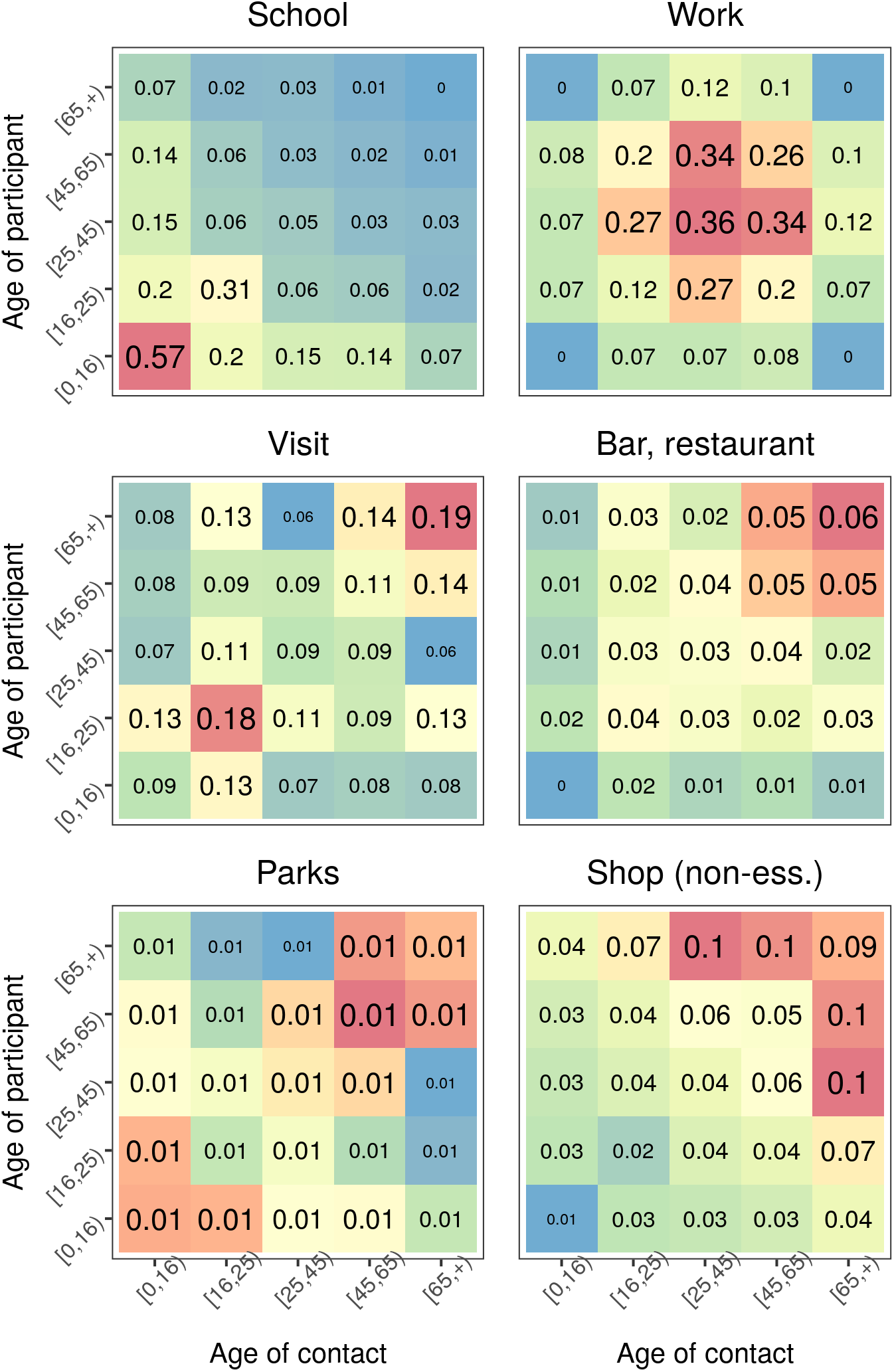
Relative change in contacts when stopping different activities. Contact rates are based on the POLYMOD contact study. Here *q_al_* is assumed to be 1.

When using the derived contact matrices in the epidemiological model for COVID-19, results were consistent in that the three activities that had the largest impact on reducing the epidemic curve and delaying its peak were school, work, and social visits (Figure 2 and Figure 3).

**Figure 2:**
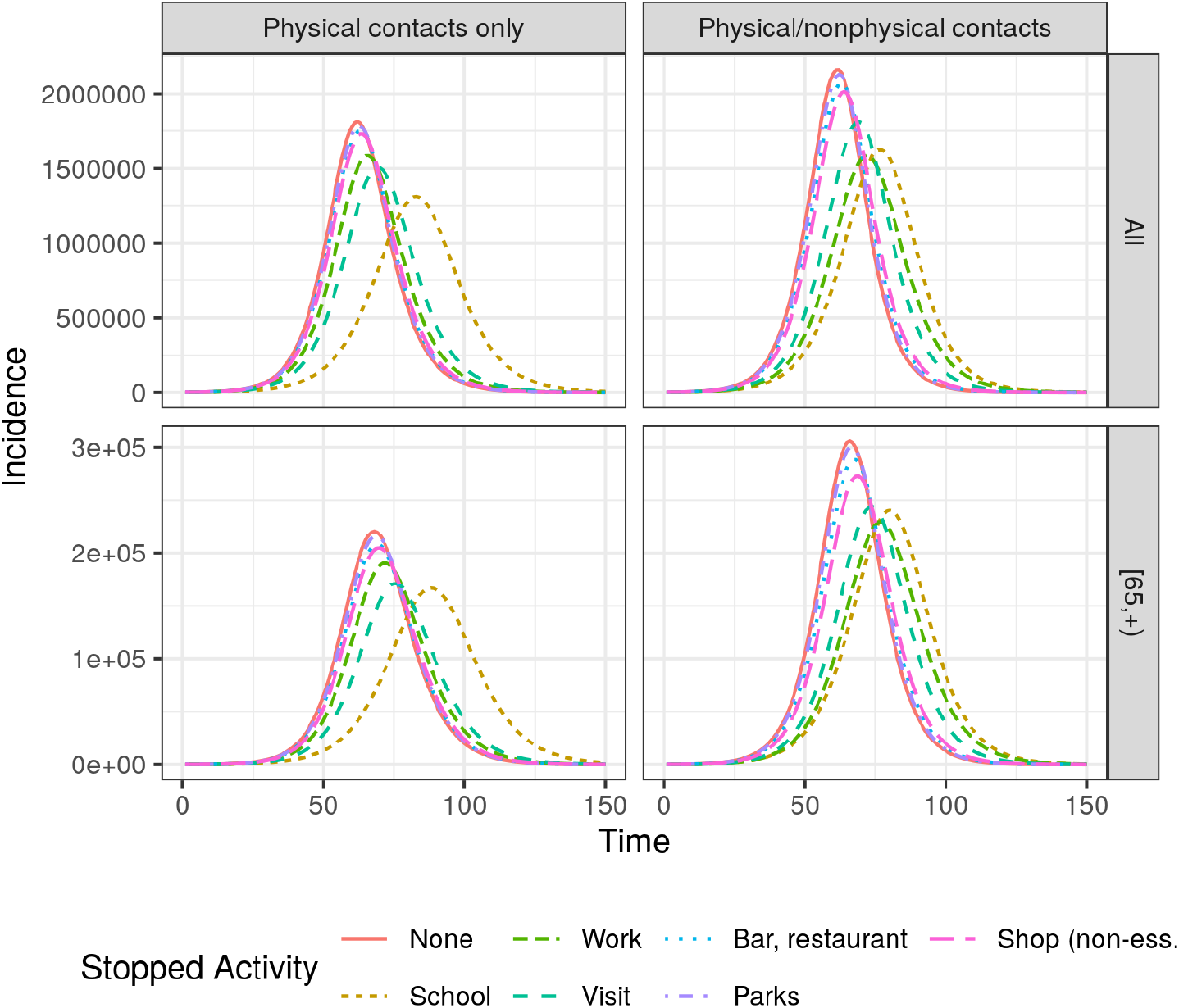
The resulting transmission dynamics when disallowing one activitiy while allowing all others to continue. Right column shows the results using physical contacts only in POLYMOD. Both the result for the whole population (top) and the elderly (bottom) is shown.

The exact impact of each activity depended on whether we used physical contacts only (right columns) or all contacts (Figure 2 and Figure 3). For physical contacts, social visits had a larger impact than work, especially in the elderly population. In the elderly the reduction in peak height is similar between stopping visits and closing schools, but closing schools would slow down the epidemic more. Looking at all contacts, work had the largest impact on peak height. This difference is probably because contacts at work were more likely to be conversational only, especially when compared to social visits and school visits. Parks and non-essential shopping had the least impact, but the impact was still significant as illustrated best when re-allowing only those activities (Figure 3)

**Figure 3:**
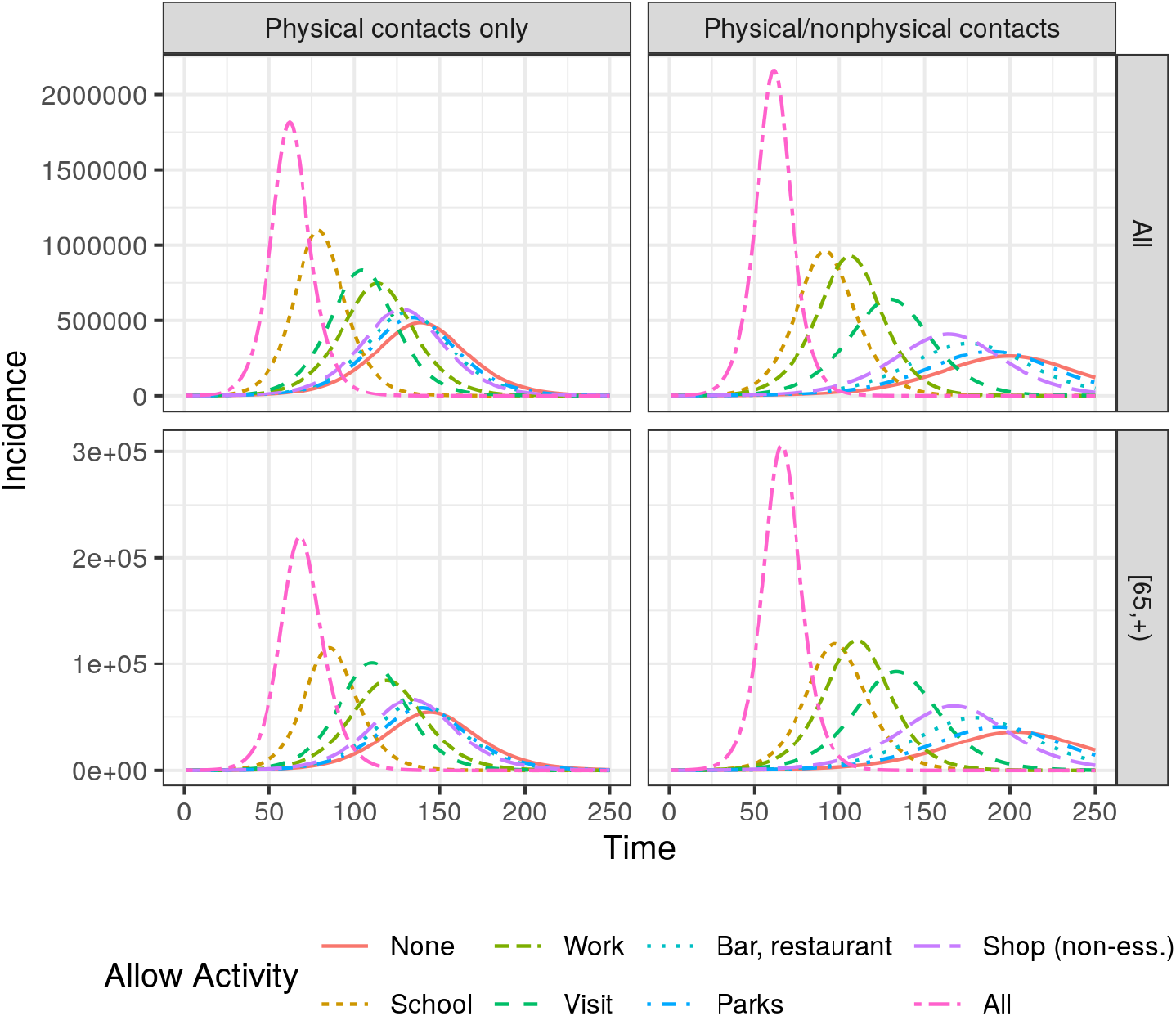
The resulting transmission dynamics when allowing one activitiy to continue while disallowing all others. Right column shows the results using physical contacts only in POLYMOD. Both the result for the whole population (top) and the elderly (bottom) is shown.

## Discussion

Social distancing is an important public health intervention to reduce or interrupt the sustained transmission of emerging infectious pathogens, for which close contact and the social contact mixing patterns of communities drives the spread [2,18,19]. The COVID-19 pandemic highlighted the need for a more fine-grained modelling of contact patterns to capture the impact of reducing or stopping selected daily activities possibly involving non-essential physical human contact.

Our findings for the augmented contact matrices showed the highest relative effect on reducing physical contacts for children and young adults aged 0-24 years when spending less time at schools/institutes of higher learning, which also still reduces contacts in adults (who may take younger children to school). For adults aged 25-64 years, closing workplaces has by far the highest effect on reducing physical contacts. For the elderly aged 65+ years, reducing the number of social visits has the highest impact on reduced contacts, with a comparable reduction also in adolescents and young adults (aged 16-24 years). Reductions of contacts were largely assortative by age, except for non-essential shopping where reductions are slightly higher between ages 25-64 years and those aged 65+ years. These findings mirror in part the underlying contact matrices, which were also highly assortative by age [13].

The dynamic-transmission model results suggest a higher fraction of close-contact transmission at workplaces, schools, and social visits than from visits to parks, bars/cafes/restaurants, and non-essential shopping. The largest impact on reducing the epidemic curve and delaying its peak was achieved when implementing all social distancing measures, followed by reducing work, schooling, and social visits. When looking at the subset of physical contacts only, the order changes to school, social visits, and working; possibly due to less physical contact and shorter duration of contacts while working. These results do depend on the assumption that the number of contacts in each activity is proportional to the time spent on that activity. This assumption might well be violated for certain activities, such as going to bars/cafes or restaurants, where the number of contacts per time spent, could be significantly higher than in other leisure activities. If more fine grained data was available then the method presented here could accommodate such concerns, by changing the activity specific weight.

These findings are in line with the modelled benefits from workplace distancing and school closures in the early stages of COVID-19 in Singapore and China [6,20]. Studies of previous pandemics also found that e.g. the early, sustained and layered application of school closures and cancellation of public gatherings during the 1918-1919 influenza pandemic were significantly associated with reductions in weekly excess deaths and delays in reaching peak mortality in the USA [21]. Obviously not all factors of influenza epidemics, particularly from over a century ago, will be present or relevant in the currently ongoing coronavirus pandemic (most notably a certain degree of pre-existing immunity in older adults and the availability of anti-virals and vaccines [22,23]).

During the previous coronavirus (SARS-CoV) pandemic in 2003 extensive measures of social distancing were also implemented in areas with widespread (suspected) community transmission, including closing schools, theatres, and public facilities as well as cancelling public mass gatherings. The combined effect led to dramatic declines in new SARS-CoV cases [18]. Nonetheless, the isolated impact of these measures remains unclear as they were adopted simultaneously, and combined with other measures of enhanced contact tracing, increased hygiene, and requiring face masks for individuals using public transport, working in restaurants, or entering healthcare facilities [18]. Moreover, the 2003 SARS-CoV outbreak may differ epidemiologically from the 2019 SARS-CoV-2 pandemic due to e.g. no pre-symptomatic transmission having been observed for the 2003 strain emergence [18], while a potential issue with SARS-CoV-2 [24]. Furthermore, transmission of the 2003 SARS-CoV strain was primarily in healthcare or household settings [25], which are characterised by close person-to-person contact [18], while the 2019 SARS-CoV-2 pandemic is characterised by widespread community transmission in a growing number of countries [3]. Taken together, the empirical evidence for the effectiveness in reducing transmission of non-pharmaceutical interventions is heterogeneous [26-28].

Our findings that the predicted impact of fewer contacts in parks, bars/cafes/restaurants, and non-essential shopping is small is suggestive of excluding these activities from the list of social distancing measures. However, reducing these activities may nonetheless be required if otherwise e.g. healthcare capacity is exceeded [29], where every little impact helps ease the pressure on services. It may thus be in the best interest of the public to not completely lift social distancing measures too early as that may inadvertently initiate an immediate second epidemic wave as seen e.g. in the 1957 influenza pandemic when schools in the Northern Hemisphere were re-opened after the summer holidays [19].

Furthermore, we were unable to account for substitution effects in the time use and activities of individuals when reducing or stopping one activity and turning instead to another, which could possibly lead to increased numbers of contacts. Previous studies on school closures found e.g. shifting mixing patterns of children to other, non-school settings, and highlighted the need for physical distancing as much as possible [30,31]. At high rates of compliance, however, the derived matrices would not need to be adjusted further. Nonetheless, mixing patterns and time-use may change immediately after re-allowing social activities for a short time, which may pose increased transmission risks especially in crowded environments indoors that may not be COVID-19-secure and if individuals chose to spend longer durations than usual at leisure activities such as bar visits. A related concern is the potentially impaired judgment of individuals following alcohol consumption. One crucial aspect to achieving widespread compliance with social distancing measures is the risk perception and the risk communication; individuals who understand the relationship of the transmission risk from direct physical contact with infectious cases who may present asymptomatic seem more likely to reduce unnecessary contacts [31-33].

### Strengths and Limitations

Our analysis combined the empirical data of two large-scale, representative surveys, and we successfully applied the resulting contact matrices in a dynamic-transmission model to explore the impact of social distancing measures adopted during the COVID-19 pandemic [3,11,12]. Augmenting the social contact matrices with time-use data has enriched the frequency of contacts with the duration of exposure in the derived matrices, which allows for more fine-grained mixing patterns than conventionally used [6,13,34]. These findings from the augmented contact matrices will be more broadly applicable to newly emerging infectious pathogens whose spread is highly dependent on the social contact mixing patterns of communities, including influenza pandemics. Our application to infections with SARS-CoV-2 causing COVID-19 illustrated the impact of reducing social contacts, and when lifting these restrictions again. Future pandemics, however, will require suitably adapted models that are tailored to the epidemiology of that pandemic pathogen and the disease it causes.

Although additional information is elicited in both surveys, we considered them the most robust for their main purpose: social contacts and the time-use per day. Furthermore, a significant amount of transport is linked in the data to school and work activities, and reductions will thus likewise impact transportation. We cannot rule out that some activities were misclassified, however; for instance, the number of contacts in bars may relate just to the group of individuals with whom someone went there instead of all individuals in close physical proximity. Also, both datasets grouped data under unspecified locations and activities that could not be used directly (about 16,000 contacts, i.e. 16% of all contacts, occurred in other places in POLYMOD; while 0.6-0.8 hours per day were unspecified in UKTUS when using our list of coding; see Table S1). Given the likely inverse relationship of the number and intensity of contacts [35], however, the impact on results may be minimal. Moreover, without knowing the amount of contacts a particular worker has, we cannot account for some complicated interactions; e.g. if people go less often to the shop then shop employees will also meet fewer people (for as long as the shops stay open altogether). We also assumed a proportional reduction in the risk of infection by number of contacts, which is a frequently made assumption but not necessarily true [35]. Moreover, transmission risks may be reduced when contacts keep a larger physical distance. All of these considerations point towards the importance of conducting additional empirical research into how, where, and with whom individuals spent their time during pandemics, and the behavioural changes that may be expected in mixing patterns as a consequence of social distancing measures (when adopted and after lifting restrictions again).

We also focused on contact matrices from empirical surveys as the predominant method emerging of parameterising social contacts in public health intervention modelling in at least a decade [5]. Although empirical diary data may suffer from recall bias and reporting inaccuracies, all other methods have their own limitations [2]. Similarly, using the data from POLYMOD and the UKTUS is illustrative of using social contact matrices and time-use data in general, which are widely used and available internationally, and largely harmonised [7], allowing to repeat our methods in different settings. Furthermore, others have successfully applied time-use data in epidemiological modelling [8-10], or reduced contact matrices by scaling factors to simulate behavioural changes in mixing [36]. Our study improves our understanding of how time-use data and mixing patterns can be combined to understand when and where transmission occurs, and which activities are to be targeted in response to social distancing measures such as adopted in many countries for the COVID-19 pandemic [3,11,12].

Our study was concerned with social distancing measures adopted domestically in populations with widespread community transmission; hence, we ignored measures such as increased hygiene and personal protective equipment, case findings approaches, or international flight restrictions and border checks [34,37,38]. Furthermore, because of the underlying data we used were gathered from members of the public in the community [13,14], we are unable to address the social mixing with (and within) other, semi-enclosed settings such as healthcare and social care settings (hospitals, care homes), military, or correctional facilities.

## Data Availability

All data has been published before.

## Acknowledgements

We gratefully acknowledge the access to the data from the United Kingdom Time Use Survey through the UK Data Service (http://doi.org/10.5255/UKDA-SN-8128-1).

## Notes

### Competing Interest Statement

The authors have declared no competing interest.

### Funding Statement

No external funding was received for this work.

